# Detection of choroidal hypoperfusion in giant cell arteritis using swept-source optical coherence tomographic angiography

**DOI:** 10.1101/2021.05.21.21257605

**Authors:** Edward S. Lu, Amy Yuan, Devon A. Cohen, Raviv Katz, John B. Miller, Eric D. Gaier

**Affiliations:** Department of Ophthalmology, Massachusetts Eye and Ear Infirmary, Boston, MA; Department of Ophthalmology, Harvard Medical School, Boston, MA; Harvard Retinal Imaging Lab; Department of Ophthalmology, University of Washington School of Medicine, Seattle, WA; Department of Ophthalmology, Boston Children’s Hospital, Boston, MA; Picower Institute for Learning and Memory, Department of Brain and Cognitive Sciences, Massachusetts Institute of Technology, Cambridge, MA

**Keywords:** ischemic optic neuropathy, giant cell arteritis, optical coherence tomographic angiography, fluorescein angiography

## Abstract

**Objective:** To determine whether swept-source optical coherence tomographic angiography (SS-OCTA) can demonstrate choroidal perfusion abnormalities seen on fluorescein angiography (FA) in giant cell arteritis (GCA).

**Design:** Observational case series.

**Participants:** Six eyes of 3 patients with bilateral ischemic optic neuropathy secondary to GCA, and one control patient without ocular involvement from biopsy-confirmed GCA.

**Methods:** *En face* SS-OCTA (DRI OCT Triton, Topcon, Tokyo, Japan) and FA centered on the macula were obtained at presentation. SS-OCTA was segmented into superficial and deep retinal capillary plexuses and the choriocapillaris laminae. SS-OCTA images were independently analyzed for perfusion abnormalities and compared with corresponding FA images.

**Main Outcome Measures:** Correspondence of choroidal angiographic abnormalities on SS-OCTA and FA.

**Results:** SS-OCTA showed decreased angiographic signal within the choriocapillaris in 5/6 eyes and corresponded to hypoperfusion abnormalities on FA in similar geographic distributions in 5/5 eyes. SS-OCTA also showed dilation of the deep retinal capillary plexus overlying the area of choroidal hypoperfusion in one eye. In the one eye without angiographic signal abnormalities on SS-OCTA, no perfusion changes were noted on FA. One control patient without ocular involvement from biopsy-confirmed GCA did not show choroidal perfusion changes on SS-OCTA or FA.

**Conclusions:** This case series demonstrates comparability between SS-OCTA and FA in detection and characterization of choroidal hypoperfusion secondary to GCA. As a rapid and non-invasive tool, SS-OCTA may serve as a viable alternative to FA in the diagnostic evaluation of GCA.

## INTRODUCTION

Giant cell arteritis (GCA) is an immune-mediated vasculitis of medium to large sized arteries that predominantly affects adults over the age of 50 with high morbidity and mortality if untreated.^1,2^ GCA affects the short posterior ciliary arteries supplying the prelaminar and laminar portions of the optic nerve head, resulting in acute vision loss and pallid nerve oedema characteristic of arteritic anterior ischemic optic neuropathy (AAION).^3,4^ Other hallmark symptoms of GCA include headache, myalgias, fatigue, fever, weight loss, or jaw claudication. Fluorescein angiography (FA) classically demonstrates delayed or incomplete choroidal filling in the acute phase of GCA.^5,6^

Optical coherence tomographic angiography (OCTA), a non-invasive tool for imaging of laminar blood flow, provides high-resolution, three-dimensional segmentation of the chorioretinal microvasculature into the superficial capillary plexus (SCP), deep capillary plexus (DCP) and choriocapillaris (CC).^7^ While FA has historically been the standard imaging modality for chorioretinal vascular pathology, OCTA is a promising alternative to dye-based angiography.^8^ OCTA has been shown to demonstrate choroidal changes in a range of eye conditions including age-related macular degeneration (AMD), central serous chorioretinopathy (CSC), uveitis, and inherited retinal disorders.^9^ Reported OCTA evidence of choroidal hypoperfusion secondary to GCA is currently limited to our previous case series and two case reports.^10–12^

We previously reported a series of 4 GCA cases imaged with spectral domain (SD)-OCTA ^10^. In 2 cases with choroidal hypoperfusion seen on FA, no corresponding reduction in SD-OCTA signal was seen. Swept-source OCTA (SS-OCTA) allows for increased penetration into the choroid compared to conventional SD-OCTA.^13–15^ We hypothesized that SS-OCTA would demonstrate choroidal perfusion abnormalities apparent on FA in patients presenting with ocular ischemia secondary to GCA.

## METHODS

### Subjects

This prospective, observational study was conducted at Massachusetts Eye and Ear and adhered to the tenets of the Declaration of Helsinki and Health Insurance Portability and Accountability Act regulations. Institutional Review Board (IRB)/Ethics Committee approval was obtained. Consent for SS-OCTA imaging and the use of imaging for education, research and publication purposes was obtained from all participants prior to enrolment. Consecutive patients with GCA who underwent imaging with both FA and SS-OCTA at the time of initial presentation between October 2017 and February 2018 were included (Table 1). GCA diagnosis was made based on a positive temporal artery biopsy or clinical features. No patients in this time interval were excluded from the study. All GCA patients had a complete dilated eye examination by an attending neuro-ophthalmologist. Automated static (Humphrey, SITA 24-2 or 30-2) or manual kinetic (Goldmann) perimetry and fundus imaging (Topcon, Tokyo, Japan) were obtained at the initial clinic visit. Demographic and clinical parameters were obtained from the electronic medical record.

**Table 1.** Patient characteristics and summary of fluorescein angiography (FA) and swept-source optical coherence tomographic angiography (SS-OCTA) findings.

| Case | Age Range (years) | Sex | ESR (mm/hr) | Laterality | Temporal artery biopsy | BCVA | DELAYED CHOROIDAL PERFUSION ON FA | DELAYED CHOROIDAL PERFUSION ON SS-OCTA |
| --- | --- | --- | --- | --- | --- | --- | --- | --- |
| 1 | 75-80 | F | 119 | OD | + | CF | Yes | Yes |
|  |  |  |  | OS | N/A | 20/30 | Yes | Yes |
| 2 | 90-95 | F | 117 | OD | + | CF* | No | No |
|  |  |  |  | OS | + | 20/30 | Yes | Yes |
| 3 | 75-80 | M | 113 | OD | N/A | 20/300 | Yes | Yes |
|  |  |  |  | OS | N/A | 20/20 | Yes | Yes |
| Control | 70-75 | F | 65 | OD | + | 20/20 | No | No |
|  |  |  |  | OS | N/A | 20/15 | No | No |

\*Baseline BCVA was CF vision from chronic macular hole.
ESR, erythrocyte sedimentation rate; BCVA, best-corrected visual acuity; FA, fluorescein angiography; SS-OCTA, swept-source optical coherence tomographic angiography; F, female; M, male; OD, right eye; OS, left eye; CF, counting fingers.

### Swept-Source OCTA

SS-OCTA images (DRI OCT Triton, Topcon, Tokyo, Japan) were obtained after pupillary dilation in the acute phase of the disease (between 1-4 days of starting corticosteroid therapy). *En face* SS-OCTA images centered on the macula (6×6 mm or 4.5×4.5 mm) were automatically segmented into the SCP (2.6 μm below the internal limiting membrane to 15.6 μm below the inner plexiform layer/inner nuclear layer interface), DCP (15.6 to 70.2 μm below the inner plexiform layer/inner nuclear layer interface) and CC lamina (0 to 10.4 μm below the basement membrane) according to the instrument’s default settings. All images were examined for quality, analyzed for perfusion abnormalities, and compared with FA. All scans were of acceptable signal quality and without significant motion artifacts.

### Fluorescein Angiography

FA images (Spectralis, Heidelberg Engineering, Heidelberg, Germany) were obtained in all cases without complication after pharmacological dilation and administration of 500 mg of IV fluorescein. Timed, choroidal filling videos and early still images (0:25-0:38) were obtained for the better-seeing eye in Case 1 and the symptomatic eyes in Cases 2 and 3. The earliest available individual FA frames were examined and are provided. Brightness and contrast were adjusted in each case to optimize visualization of perfusion defects.

## RESULTS

### Case 1

A woman in her late 70s with a history of rheumatoid arthritis presented with sudden vision loss in the right eye (OD) one day prior, with jaw claudication, headache, scalp tenderness and weight loss. On examination, visual acuities were counting fingers OD and 20/30 in the left eye (OS). There was a brisk relative afferent pupillary defect OD and a dense inferior arcuate defect OS on visual field testing (figure 1F; inset). Funduscopic exam revealed pallid optic disc oedema that was circumferential in the right eye and most prominent superonasally in the left eye (figure 1A, F). Inflammatory markers and platelets (PLT) were elevated (ESR 119 mm/hour, CRP 75 mg/L, PLT 748 K/uL). Right temporal artery biopsy was positive. FA demonstrated a large, sharply demarcated area of choroidal filling delay involving the temporal fundus in the right eye (figure 1B). The left eye also demonstrated a large, sharply demarcated area of delayed choroidal perfusion centrally that extended inferiorly and temporally (figure 1G). Late frames showed full eventual choroidal filling in these areas and leakage at the optic nerve head in both eyes (not shown). SCP and DCP segmentation of *en face* SS-OCTA images demonstrated reduced angiographic signal in the perfusion beds of small cilioretinal arteries (figures 1C-D, H-I; green arrows). There was reduction in the SCP density temporally in the right eye, potentially signifying an early consequence of ganglion cell layer and retinal nerve fiber layer atrophy. Segmentation of the CC demonstrated decreased perfusion in the same distribution as the choroidal filling delay observed on FA (figure 1E, J). Relative dilation of the overlying DCP was appreciated in the less severely affected left eye (figure 1I, yellow arrows).

**Figure 1.**
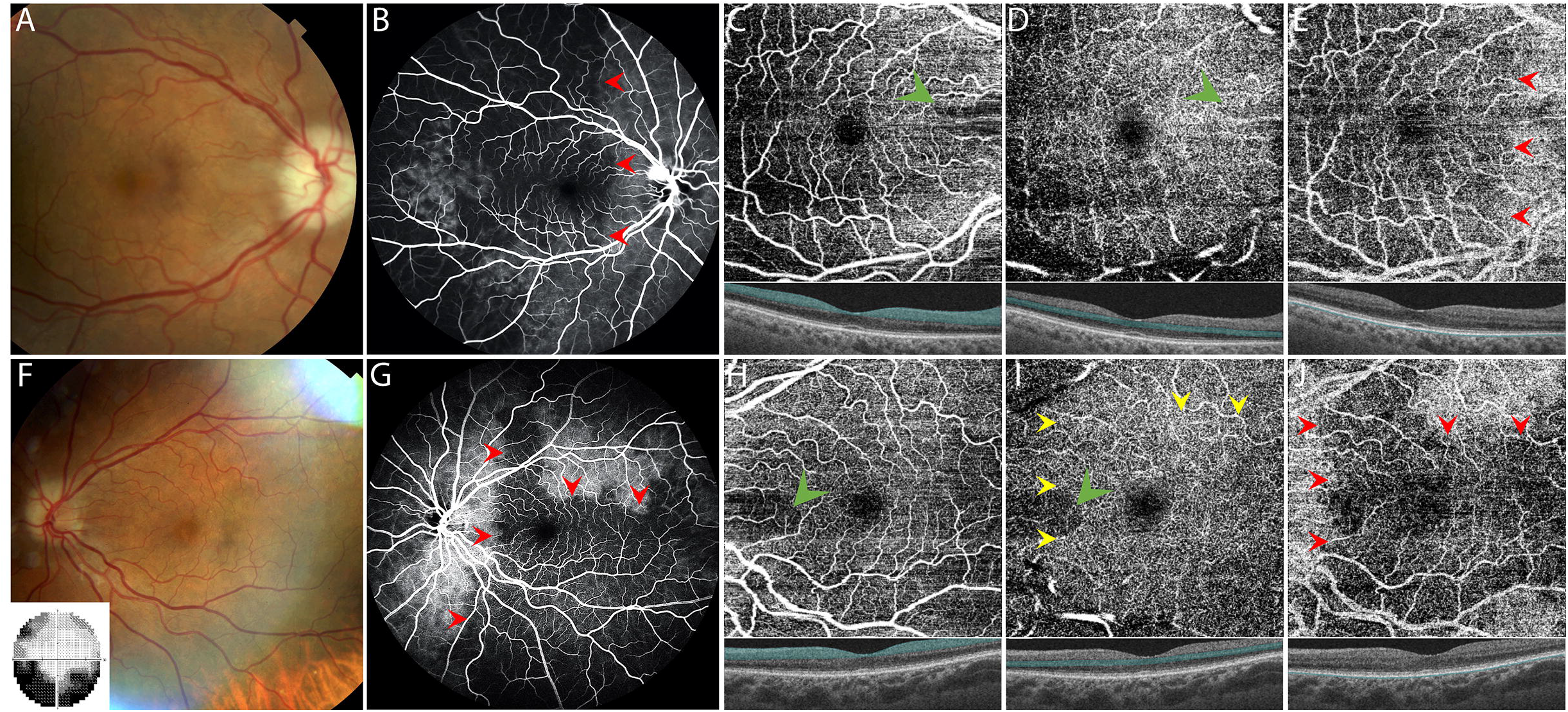
Case 1: A woman in her 70s with GCA causing sudden vision loss OD. Fundus images of the right (A) and left (F) eyes. Automated (Humphrey) perimetry (30-2) of the left eyes (F; inset). FA images taken in the mid-phase (1:21) and early phase (0:25) for the right (B) and left (G) eyes, respectively. *En face* SS-OCTA 6×6 mm images with laminar segmentation of the superficial capillary plexus (C, H), deep capillary plexus (D, I) and choriocapillaris (E, J) for the right and left eyes, respectively. Insets depict B-scan images through the fovea with segmented area highlighted in blue. Red arrows outline matching boundaries of perfusion/nonperfusion on FA and SS-OCTA. Green arrows indicate decreased angiographic signal in the perfusion beds of small cilioretinal arteries. Yellow arrows denote areas of DCP dilation.

The patient was treated with intravenous methylprednisolone and transitioned to an oral prednisone taper. Follow-up exam 6 months later revealed visual acuities of no light perception OD and improvement to 20/25 OS with stable visual field defects.

### Case 2

A woman in her 90s with a chronic macular hole in the right eye presented with one week of night sweats, headaches, scalp tenderness, and jaw claudication without subjective visual loss. Inflammatory markers were markedly elevated (ESR 117 mm/hour and CRP 298 mg/L), and bilateral temporal artery biopsies were positive. Visual acuity was unchanged from baseline (counting fingers OD due to macular hole and 20/30 OS), and a relative afferent pupillary defect OD was present. Visual field testing, notable for excessive fixation losses OS and false negatives OU, showed inferior altitudinal defects OD>OS (figure 2A, F; insets). Fundus exam was notable for bilateral pallid optic disc oedema and nasal optic disc hemorrhage in the left eye (figure 2A, F). FA demonstrated focal delayed choroidal filling in the inferior macula of the left eye (figure 2G, red arrow). SCP and DCP segmentation of SS-OCTA *en face* imaging showed foveal and perifoveal changes consistent with chronic macular hole in the right eye ^16^ (figure 2C, D) and mild nasal reductions in the SCP and DCP densities in the left eye (figure 2H, I). Although SS-OCTA imaging did not capture the full fundus view obtained on FA, CC segmentation showed an inferior region of decreased signal extending toward the fovea (figure 2J, red arrows). No choroidal filling delay in the right eye was observed on FA or SS-OCTA (figure 2B-E).

**Figure 2.**
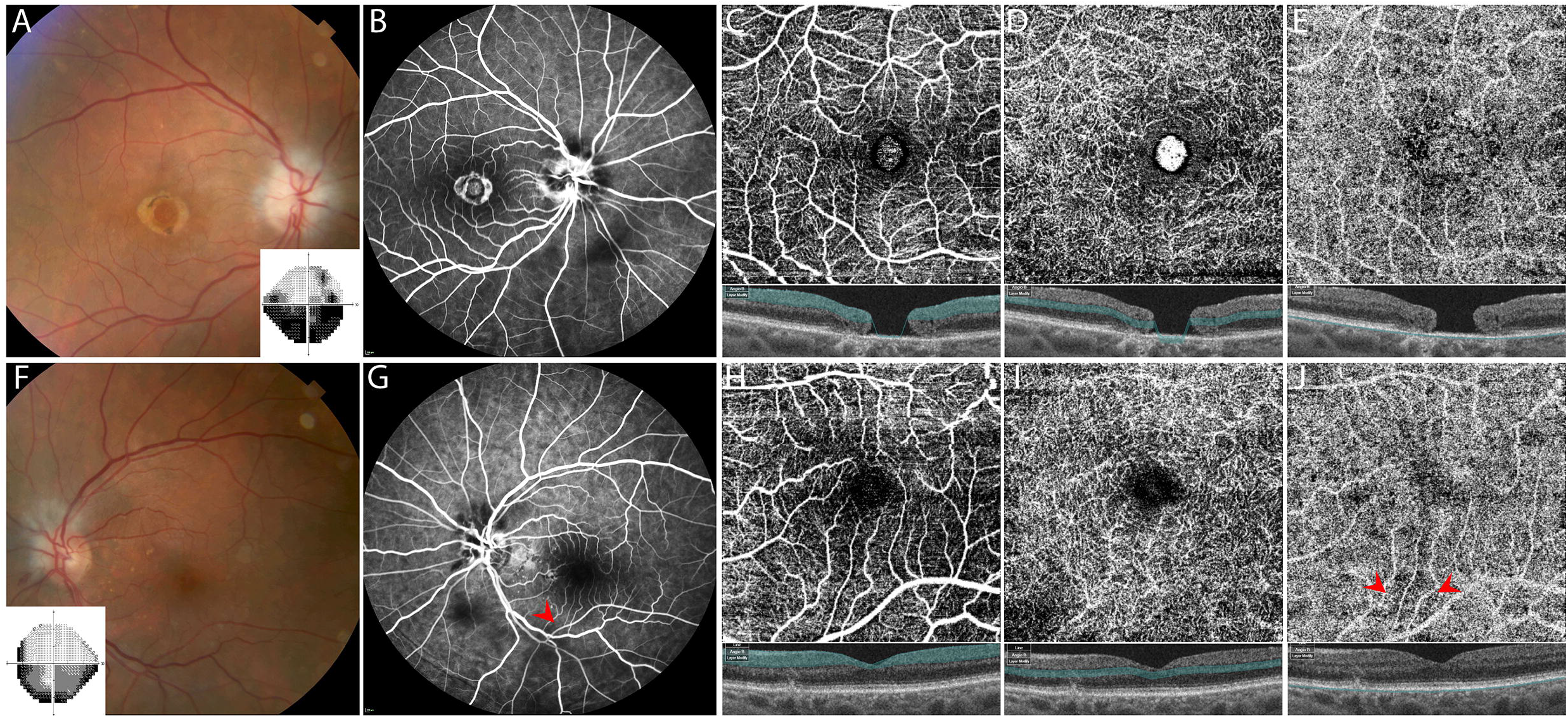
Case 2: A woman in her 90s with chronic macular hole in the right eye and systemic symptoms of GCA. Fundus images of the right (A) and left (F) optic discs. Automated (Humphrey) perimetry (24-2) of the right and left eyes (A, F; insets). FA images taken in the mid-phase (1:02) and early phase (0:38) for the right (B) and left (G) eyes, respectively. *En face* SS-OCTA 4.5×4.5 mm images with laminar segmentation of the superficial capillary plexus (C, H), deep capillary plexus (D, I) and choriocapillaris (E, J) for the right and left eyes, respectively. Insets depict B-scan images through the fovea with segmented area highlighted in blue. Red arrows indicate areas of hypoperfusion on FA and SS-OCTA. In (C) and (D), the bright signal in the centre is due to incorrect segmentation of the macular hole by the software algorithm.

The patient was started on high-dose oral prednisone with resolution of her systemic symptoms. Follow-up exam 9 months later revealed stable vision without signs of active ocular involvement of GCA.

### Case 3

A man in his late 70s with a history of polymyalgia rheumatica (PMR) presented with a 3-day history of acute, painless vision loss OD and jaw claudication. Maintenance oral prednisone for his PMR had been reduced from 20 mg to 5 mg daily three months prior. ESR was 126 mm/hr, and CRP was 6.4 mg/L. Temporal artery biopsy was not obtained due to high clinical suspicion for GCA. Exam revealed visual acuities of 20/300 OD and 20/20 OS, dyschromatopsia OD, and a relative afferent pupillary defect OD. Automated perimetry showed a dense superior paracentral defect OD and nonspecific defects OS (figure 3A, F; insets). Fundus exam was significant for inferior sectoral pallid optic disc oedema with hemorrhages in the right eye and optic disc oedema with superimposed nasal cotton wool spots in the left eye (figure 3A, F). FA showed optic nerve leakage and areas of hypofluorescence in the inferior macula likely (figure 3B, G). SS-OCTA demonstrated a reduction in SCP density in the right eye likely corresponding to inner retinal atrophy (figures 3C, H; blue arrows), and DCP dilation temporally in the left eye (figure 3D, I; yellow asterisk). In the CC, SS-OCTA showed mildly decreased signal inferior to the fovea along the inferior arcade that was apparent on FA in the right eye (figure 3B, E; red arrows). Similarly, subtle decrease in SS-OCTA CC angiographic signal inferior to the fovea on SS-OCTA corresponded to the area of hypofluorescence on FA (figure 3G, J; red arrows).

**Figure 3.**
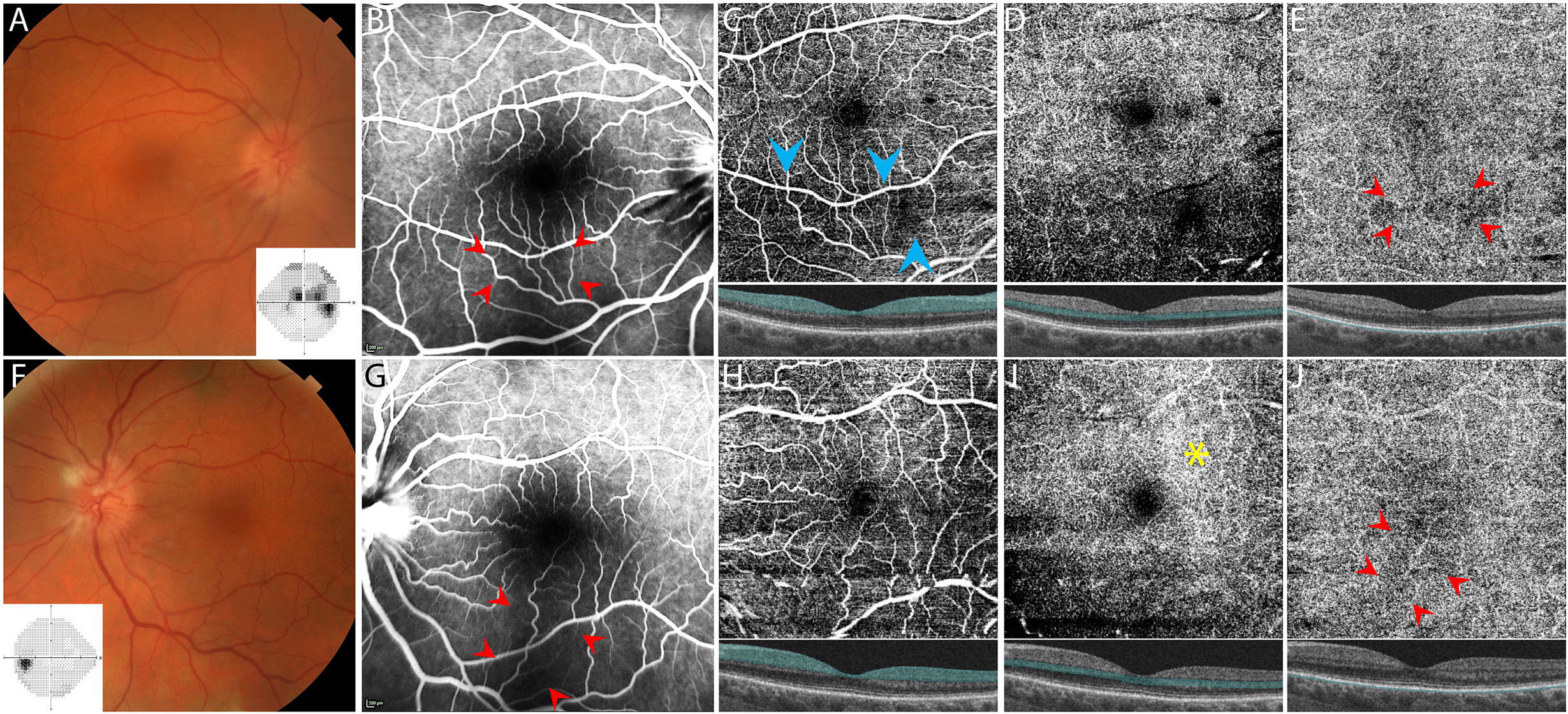
Case 3: A man in his 70s with vision loss OD. Fundus images of the right (A) and left (F) optic discs. Automated (Humphrey) perimetry (24-2) of the right and left eyes (A, F; insets). FA images taken in the mid-phase (1:09; 2:24) for the right (B) and left (G) eyes, respectively. *En face* SS-OCTA 6×6 mm images with laminar segmentation of the superficial capillary plexus (C, H), deep capillary plexus (D, I) and choriocapillaris (E, J) for the right and left eyes, respectively. Insets depict B-scan images through the fovea with segmented area highlighted in blue. Red arrows indicate areas of hypoperfusion on FA and SS-OCTA. Blue arrows indicate decreased angiographic signal on SS-OCTA likely due to inner retinal atrophy. Yellow asterisk denotes area of DCP dilation.

The patient was treated with intravenous methylprednisolone followed by an oral prednisone taper. Visual acuity improved to 20/50 OD. Two months later, the patient had a recurrence on 30 mg of prednisone, presenting with visual loss OS, prompting an increase in oral prednisone.

### GCA without choroidal hypoperfusion

One patient without visual symptoms and biopsy-proven GCA was imaged with SS-OCTA as a control (online supplementary figure 1). The patient was a woman in her 70s referred by her neurologist to evaluate for ophthalmic signs suggestive of GCA in the setting of elevated inflammatory markers (erythrocyte sedimentation rate (ESR) 65 mm/hour and C-reactive protein (CRP) 111 mg/L), jaw claudication (confounded by active dental disease), weight loss, and headaches with scalp tenderness. The patient denied visual changes. FA was obtained given her atypical clinical presentation, and no choroidal filling defects were found. Subsequent right temporal artery biopsy was positive, and the patient was treated with corticosteroids.

## DISCUSSION

Choroidal filling delay demonstrated by IV dye-based angiography is a highly suggestive finding of GCA in the right clinical context, due to hypoperfusion of the posterior ciliary arteries.^6^ As such, FA serves as a useful ancillary tool in the evaluation of GCA, particularly in mild cases. To our knowledge, this is the first clinical series applying SS-OCTA to image choroidal perfusion abnormalities in GCA. Among eyes with delayed or incomplete perfusion defects on FA, SS-OCTA showed decreased signal in the CC in all cases (5/5 eyes). Spatial mapping of choroidal hypoperfusion with SS-OCTA matched that delineated by FA with high fidelity. Among cases without choroidal filling abnormality on FA, 3/3 eyes showed no choroidal signal abnormalities on SS-OCTA. In 2 eyes with choroidal hypoperfusion, SS-OCTA showed dilation of the overlying deep retinal capillary plexus in a similar distribution (figures 1I, 3F). Thus, our results support comparability between FA and SS-OCTA to detect choroidal hypoperfusion secondary to GCA.

Prior studies using SD-OCTA have failed to show clear choroidal perfusion abnormalities that were correspondingly evident on dye-based angiography.^10,11^ We previously reported 4 GCA cases in which SD-OCTA demonstrated superficial peripapillary dilation and retinal capillary perfusion defects that corresponded to visual field loss in the acute phase.^10^ Two patients in the series had choroidal perfusion abnormalities evident on FA, yet SD-OCTA analysis of the choroid and CC (performed on the same day as FA) showed no signal abnormality in those corresponding regions. Balducci et al. reported a single case of GCA in which SD-OCTA was reported to demonstrate a sectoral peripapillary choroidal perfusion defect with correspondence on FA.^11^ However, the region in question corresponded to the perfusion bed of a cilioretinal artery, which was hypoperfused, likely edematous and imparted a blockage effect on both FA and SD-OCTA. Nemiroff et al. reported a case of GCA with decreased CC vessel density in a triangular distribution on SD-OCTA that co-localized to a large Amalric infarct on FA ^17,18^. Amalric infarcts are notably on the more severe end of the spectrum. Our present series includes cases on the milder end of the spectrum, which nonetheless manifested as decreased focal choroidal perfusion on segmentation of the CC on SS-OCTA.

Our findings support our hypothesis that SS-OCTA offers improved resolution of deeper retinal and choroidal vessels as compared to SD-OCTA and thus might be better suited to capture alterations in choroidal perfusion.^19^ Comparative studies have demonstrated improved visualization of the choroid attributed to enhanced choroidal penetration using SS-OCTA compared to SD-OCTA in eyes with clinically significant cataract and AMD.^20,21^ In addition, SS-OCTA has demonstrated greater accuracy in detecting CC flow impairment under drusen and visualizing the macula in gas-filled eyes post-vitrectomy for macular holes and retinal detachment.^22,23^ Tran et al. reported the first definitive case of choroidal hypoperfusion secondary to GCA imaged using SS-OCTA to offer a direct comparison with FA and ICG.^12^ The case depicted a large and dense, Amalric choroidal infarct that is most similar to the right eye of case 1 depicted here (figure 2A-E). Of note, the case depicted in Tran et al. employed a different SS-OCTA device (Plex-Elite 9000, Carl Zeiss Meditec, Dublin, CA) than the one used in our study and allowed for direct OCTA imaging of the full choroid. While their findings are similar to our findings in case 1, there are likely to be differences between SS-OCTA devices that influence sensitivity in detecting subretinal perfusion abnormalities.

OCTA has the potential to serve as a practical alternative to FA. There are several barriers and limitations to obtaining FA testing, including the need for a licensed practitioner to place an IV and administer fluorescein, the longer time required to complete the test, adverse reactions to fluorescein dye, %),^24^ and the inability to repeat FA testing in the event of poor image quality until the dye is excreted. On the other hand, OCTA is fast, easy to capture and non-invasive.^7^ Thus, OCTA may enable more expedient assessment of choroidal perfusion than dye-based angiography to facilitate the diagnosis and treatment of GCA in select cases.

OCTA provides higher resolution and laminar segmentation as compared with FA, allowing for the detection of lamina-specific changes that might otherwise be obscured or not appreciable on 2D FA. Segmentation of the SCP on SS-OCTA showed early reductions in vascular density that corresponded with visual defects on clinical examination and automated perimetry (figure 4C).^10^ In the DCP, we observed increased angiographic signal overlying areas of choroidal hypoperfusion in 1 of 5 GCA eyes with choroidal perfusion abnormalities. It is unclear what this increase in angiographic signal represents. Retinal vessels lack autonomic innervation and rely on autoregulation based on local factors to maintain perfusion.^25^ While choroidal perfusion changes have been implicated in retinal dysfunction in glaucoma, diabetes, hypertension, and AMD,^26^ outer retinal abnormalities associated with choroidal infarcts in GCA have been reported,^27^ but the effect of choroidal changes on the retinal capillary plexuses in acute GCA remains unclear. Specifically, further work is needed to determine the geographical correspondence between DCP and CC changes. Finally, OCTA allows for more accurate distinction of true perfusion abnormalities from artifacts (such as blockage artifacts) through assessment of signal abnormalities across lamina.^10^ Thus, laminar segmentation affords several advantages of OCTA over FA.

FA and SS-OCTA share some limitations. The absence of choroidal perfusion abnormalities does not rule out GCA regardless of the imaging modality used (e.g., see GCA control; figure 1A-J). Choroidal hypoperfusion, whether captured on FA or SS-OCTA, is not specific to GCA and has been reported in AMD, CSC, diabetic retinopathy, acute posterior multifocal placoid pigment epitheliopathy, and retinitis pigmentosa.^28–32^ As is the case with FA, choroidal perfusion abnormalities suggested by OCTA must be interpreted as part of a complete clinical evaluation for GCA. Ambiguous OCTA findings could be corroborated by FA and vice versa.

Our study is limited by a relatively small number of total affected eyes. However, it is worth noting that each case can be considered separately since within-subject comparisons demonstrate remarkable spatial fidelity across a range of hypoperfusion severities. Our study approach was qualitative, and therefore identification and assessment of signal abnormalities on SS-OCTA may be subject to confirmation bias, just as they are in ambiguous cases imaged by FA. Unlike FA, SS-OCTA is amenable to automated quantification that may increase the sensitivity to detect more subtle perfusion abnormalities. Further study and efforts to automate detection of angiographic signal changes in ischemic neuro-ophthalmic disease would be of great benefit. Finally, we could not make a direct comparison between SD-OCTA and SS-OCTA because GCA cases presenting to our service were imaged with one modality or the other. However, cases showing areas of hypoperfusion were comparable in size and density between our prior study ^10^ and this study, thus supporting greater sensitivity of SS-OCTA.

In conclusion, we demonstrate high comparability between SS-OCTA and FA in detecting choroidal ischemia in GCA. There are many advantages of SS-OCTA over FA, including ease and expedience of obtaining imaging in the acute setting that is highly relevant in clinical scenarios of suspected GCA. Further study is needed to evaluate SS-OCTA as a diagnostic tool in screening for choroidal perfusion abnormalities when GCA is suspected.

## Data Availability

Raw data are available upon reasonable request to the corresponding author.

## Abbreviations

(SS-OCTA): swept-source optical coherence tomographic angiography
(FA): fluorescein angiography
(GCA): giant cell arteritis
(AAION): arteritic anterior ischemic optic neuropathy
(SCP): superficial capillary plexus
(DCP): deep capillary plexus
(CC): choriocapillaris
(AMD): age-related macular degeneration
(CSC): central serous chorioretinopathy
(SD): spectral-domain
(PLT): platelet
(ESR): erythrocyte sedimentation rate
(PMR): polymyalgia rheumatica
(CRP): C-reactive protein

## FIGURE LEGENDS

**Online supplementary figure 1**. Swept-source optical coherence tomographic angiography (SS-OCTA) images from a control, GCA-positive patient in her 70s without ocular involvement. Fundus images of the right (A) and left (F) eyes. Fluorescein angiography (FA) images taken in the early phase (0:17) and mid-phase (0:46) for the right (B) and left (G) eyes, respectively. *En face* SS-OCTA 6×6 mm images with laminar segmentation of the superficial capillary plexus (C, H), deep capillary plexus (D, I) and choriocapillaris (E, J) for the right and left eyes, respectively. Horizontal lines on SS-OCTA images are due to motion artifact.

