## Supplementary figures and images for "Detection of choroidal hypoperfusion in giant cell arteritis using swept-source optical coherence tomographic angiography"

### Supplemental Figure 1

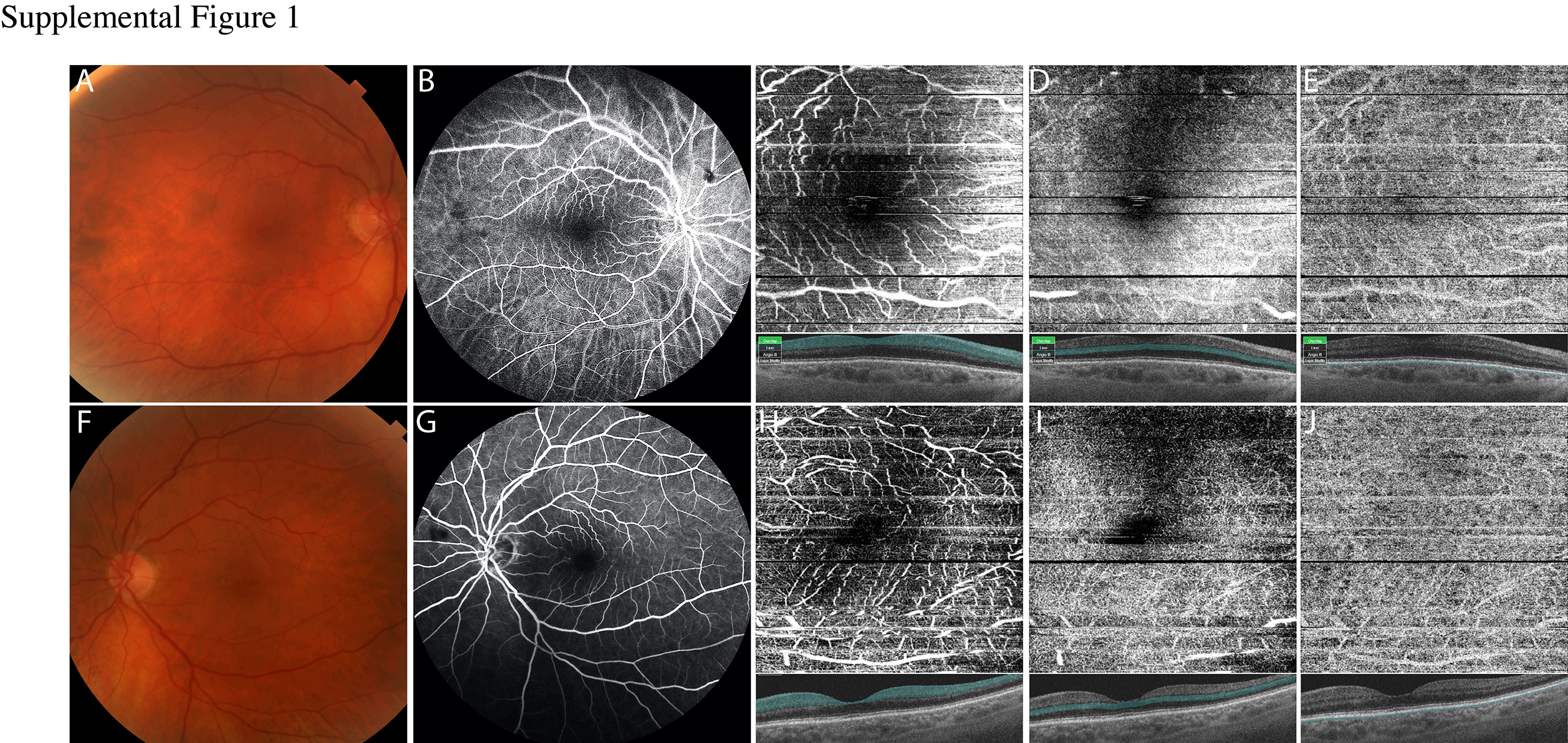
